# Association of non-HDL-C/apoB ratio with long-term mortality in the general population: a cohort study

**DOI:** 10.1101/2023.10.17.23297181

**Authors:** Kerui Zhang, Chenchen Wei, Yaqing Shao, Li Wang, Zongquan Zhao, Song Yin, Xuejun Tang, Yuan Li, Zhongshan Gou

## Abstract

**Background:** In general, the identification of cholesterol-depleted lipid particles can be inferred from non-high-density lipoprotein cholesterol (non-HDL-C) concentration to apolipoprotein B (apoB) concentration ratio, which serves as a reliable indicator for assessing the risk of cardiovascular disease. However, the ability of non-HDL-C/apoB ratio to predict the risk of long-term mortality among the general population remains uncertain. The objective of this study is to explore the association of non-HDL-C/apoB ratio with long-term all-cause and cardiovascular mortality in the adult population of the United States.

**Methods:** This retrospective cohort study was a further analysis of existing information from the National Health and Nutrition Examination Survey (NHANES). In the ultimate analysis, 12,697 participants from 2005 to 2014 were included. Kaplan-Meier (K-M) curves and the log-rank test were applied to visualize survival differences between groups. Multivariate Cox regression and restricted cubic spline (RCS) models were applied to evaluate the association of non-HDL-C/apoB ratio with all-cause and cardiovascular mortality. Subgroup analysis was conducted for the variables of age, sex, presence of coronary artery disease, diabetes and hypertriglyceridemia and usage of lipid-lowering drugs.

**Results:** The average age of the cohort was 46.8 ± 18.6 years, with 6,215 (48.9%) participants being male. During a median follow-up lasting 68.0 months, 891 (7.0%) deaths were documented and 156 (1.2%) patients died of cardiovascular disease. Individuals who experienced all-cause and cardiovascular deaths had a lower non-HDL-C/apoB ratio compared with those without events (1.45 ± 0.16 *vs.* 1.50 ± 0.17 and 1.43 ± 0.17 *vs.* 1.50 ± 0.17, both *P* < 0.001). The results of adjusted Cox regression models revealed that non-HDL-C/apoB ratio exhibited independent significance as a risk factor for both long-term all-cause mortality [hazard ratio (HR) = 0.51, 95% confidence interval (CI): 0.33-0.80] and cardiovascular mortality (HR = 0.33, 95% CI: 0.12-0.90). Additionally, a significant sex interaction was discovered (*P* for interaction < 0.05), indicating a robust association between non-HDL-C/apoB ratio and long-term mortality among females. The RCS curve showed that non-HDL-C/apoB ratio had a negative linear association with long-term all-cause and cardiovascular mortality (*P* for non-linearity was 0.098 and 0.314).

**Conclusions:** The non-HDL-C/apoB ratio may serve as a potential biomarker for predicting long-term mortality among the general population, independent of traditional risk factors.

## Introduction

The association of abnormal lipid metabolism and atherosclerotic cardiovascular disease (ASCVD) has been extensively established, and ASCVD has become the foremost global cause of mortality[1, 2]. Non-high-density lipoprotein cholesterol (non-HDL-C) contains all the cholesterol present in lipoprotein particles resulting in ASCVD, could explain the residual risk not attributable to low-density lipoprotein cholesterol (LDL-C) or other established risk factors[3]. Apolipoprotein B (apoB) is the primary apoprotein in non-HDL-C[4], can be used to measure the amount of atherogenic lipoprotein particles present in blood[5]. Still, non–HDL-C is unable to characterize dyslipidemias[6] and differentiate between remnant cholesterol (RC) and LDL-C[7]. Discordance analysis reveals that the numbers of atherogenic particles are the primary determinant of risks of ASCVD, especially when numbers and concentration are inconsistent[8–10]. The non-HDL-C/apoB ratio serves as a simple physiologically relevant ratio to reflect this inconsistency and distinguish the relative amounts of cholesterol-depleted and cholesterol-enriched particles[11]. Nowadays, nuclear magnetic resonance (NMR) spectroscopy is a reliable tool for determining the lipoprotein profile, with outcomes that are generally unaffected by the composition of lipoproteins. Nevertheless, pricey equipment and strict testing requirements make it challenging to promote[12, 13]. Ion mobility analysis and vertical auto profile are both viable alternatives as well[14]. Considering the complexity of above-mentioned testing technologies, non-HDL-C/apoB ratio could function as a more readily available and cost-effective tool to assess numbers of atherogenic particles especially cholesterol-depleted particles.

Previous studies[15, 16] on the role of non-HDL-C/apoB ratio in diseases mostly concentrated on the simplified diagnosis algorithm for type III hyperlipoproteinemia (familial dysbetalipoproteinemia). A study of South Asian individuals reported that non-HDL-C/apoB ratio < 1.4 will markedly increase coronary calcification at any age[11]. It is worth mentioning that similar researches mainly focus on LDL-C/apoB ratio. This indicator serves as a measurement of LDL particle size and can be used to predict the occurrence of cardiovascular events and deaths[17–19]. However, measurements of LDL-C, whether indirect estimation methods or direct methods such as the β-quantification procedure and precipitation method, are susceptible to high triglyceride (TG) level, leading to inaccurate results[20, 21]. Furthermore, LDL-C underestimates the concentration of other pathogenic particles in non-HDL-C. Given the above factors, non-HDL-C/apoB ratio may be a better method for evaluating cardiovascular risks. To the best of our knowledge, there are no studies that focus on non-HDL-C/apoB ratio to predict long-term mortality in the general population. Consequently, the primary aim of our study is to explore the association between non-HDL-C/apoB ratio and long-term mortality among individuals in general.

## Methods

### Study design and cohort

The study cohort was obtained from the official website of the National Health and Nutrition Examination Survey (NHANES), a series of complex surveys designed to capture data from the non-institutionalized civilian population in the United States. The comprehensive protocol can be found in the NHANES process manual (https:// www.n.cdc.gov/nchs/nhanes/analyticguidelines.aspx). The health survey encompassed a cohort of 50,965 individuals from 2005 to 2014. All individuals with available data on total cholesterol (TC), HDL-C and apoB were included in this study (n=15,697). There were 3, 617 cases who were excluded from this study, of which 3, 000 cases were under the age of 18, 604 cases had cancer at baseline, and 13 cases missed follow-up. And the remaining 12, 697 participants were enrolled for further analysis (**Figure 1**). The program passed ethical review, and all participants provided informed consent.

**Figure 1.**
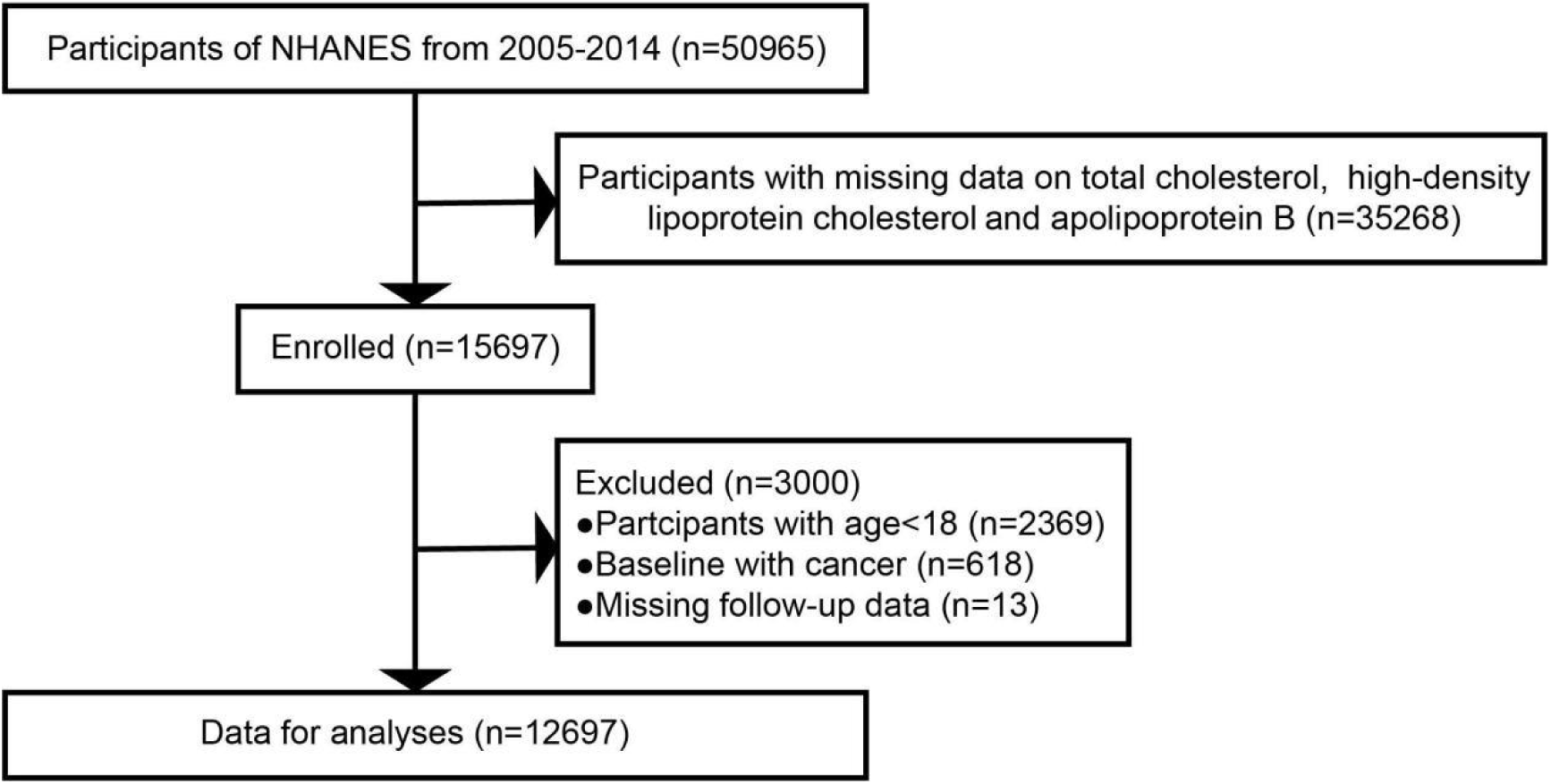
Flow diagram of study selection. NHANES, National Health and Nutrition Examination Survey.

### Assessment of exposure

Fasting blood samples were collected in accordance with established venipuncture protocol and procedure. The quantification of TC and TG values was measured enzymatically[22]. Serum HDL-C value was determined by precipitation method on the Roche Modular P and Roche Cobas 6000 chemistry analyzers, while apoB was measured by immunonephelometry[19]. Non-HDL-C was calculated by subtracting HDL-C from TC.

### Covariates

Study cohort were interviewed face-to-face or via computer-assisted personal interview. Participants filled out a questionnaire with demographic and health details. Demographic information including date of birth, sex, racial background, health-related details (such as medical history and habits related to smoking and alcohol consumption), and the medication at baseline, was collected. Based on self-reported information, data regarding the history of medical diseases: hypertension, diabetes and coronary artery disease (CAD) was gathered too. Subjects who had duration smoking for exceeding 6 months or over 100 cigarettes were considered smokers, and subjects who had drank at least 12 drinks in the last year were considered drinkers[23]. Body mass index (BMI) was calculated as weight (kg)/height squared (m^2^). Participants who had systolic blood pressure ≥ 140 mmHg and/or diastolic blood pressure ≥ 90 mmHg, a history of hypertension, or were taking antihypertensive drugs were classified as having hypertension[24]. Participants with fasting blood glucose value ≥ 7 mmol/L or glycated hemoglobin value ≥ 6.5% were classified as having diabetes[25].

### Ascertainment of outcomes

The causes of death were categorized based on the 10th edition of the International Classification of Diseases (ICD-10). The endpoints were all-cause and cardiovascular mortality (defined as cardiovascular disease-related death (codes I00-I99)). The collection of mortality data was accomplished through cross-referencing the NHANES datasets with the National Death Index (https://www.cdc.gov/nchs/data-linkage/mortality.htm). The follow-up period lasted from the date of survey participation to death, the date of drop-out, or December 31, 2015, whichever event transpired first.

### Statistical analysis

Continuous variables were represented using the mean ± standard deviation and categorical variables were expressed as numbers with percentages. The Mann-Whitney U or chi-square tests were employed for group comparisons. The study employed Kaplan-Meier (K-M) curves and log-rank tests to assess and evaluate survival differences between groups. The multiple Cox regression models were built to explore the independent association between non-HDL-C/apoB ratio and long-term mortality. Model 1 was a crude model without confounder adjustments. Model 2 incorporated covariates such as age, sex, race, drinking and smoking status. Model 3 incorporated all covariates present in Model 2, alongside additional conventional cardiovascular risk factors, including hypertension, diabetes, CAD, TG value, and lipid-lowering drugs use.

To conduct subgroup analysis, we examined the results stratified by age, sex, use of lipid-lowering drugs, and presence of diabetes, CAD or hypertriglyceridemia (defined as serum TG level > 1.7 mmol/L) from the fully adjusted models. Interactions among these variables were also evaluated. Additionally, we assessed the linear trend between non-HDL-C/apoB ratio and long-term mortality risk using restricted cubic spline (RCS) models. Missing values were replaced using multiple imputation. Statistical analyses were conducted using R version 4.2.0, employing two-tailed testing. The value of *P* < 0.05 was regarded as statistically significant.

## Results

### Baseline characteristics

The mean age of the entire study cohort (n = 12,697) was 46.8 ± 18.6 years, and 6,215 (48.9%) were male. The average non-HDL-C/apoB ratio was 1.50 ± 0.17. The percentile spread from 1.26 to 1.74 (5th to 95th percentile) and from 1.15 to 1.96 (1st to 99th percentile). Over a median follow-up period of 68.0 months, 891 (7.0%) all-cause deaths occurred, 156 (17.5%) of which were attributed to cardiovascular disease.

The study cohort was categorized into two groups based on long-term all-cause mortality: subjects without events (n = 11,806) and with events (n = 891). A comparison of the demographic and baseline characteristics of the two groups was presented in **Table 1**. Participants in group with events were older, mostly male, and had a larger percentage of smokers and drinkers (all *P* values < 0.05). Any cardio-cerebrovascular diseases, such as hypertension, diabetes, CAD, were more common in the population experiencing all-cause mortality, as was the use of lipid-lowering drugs (all *P* values < 0.05). Moreover, they tended to have higher serum TC, TG, LDL-C and non-HDL-C concentrations (all *P* values < 0.05). Importantly, participants with events had significantly lower non-HDL-C/apoB ratio (*P* < 0.001).

**Table 1.**
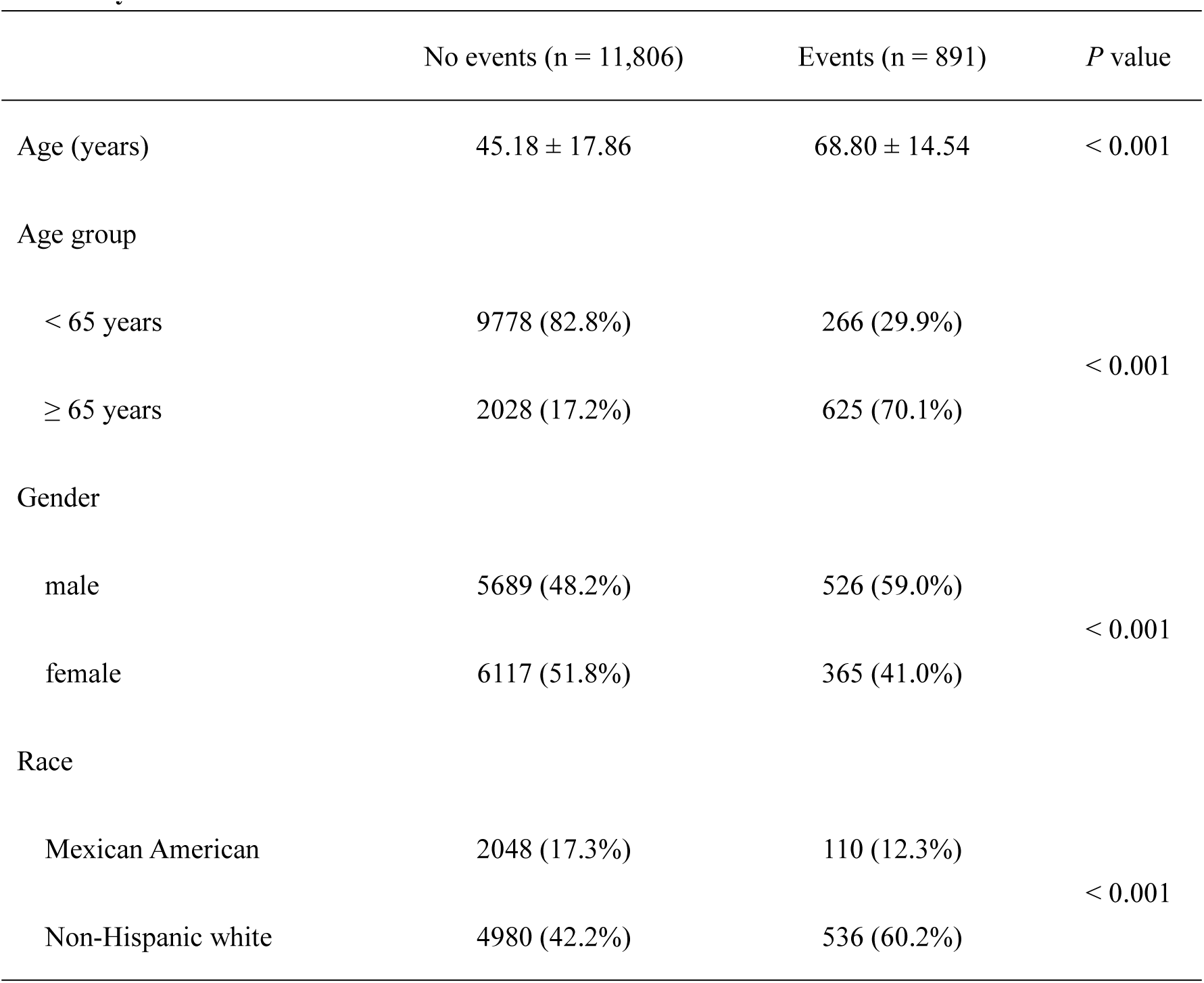
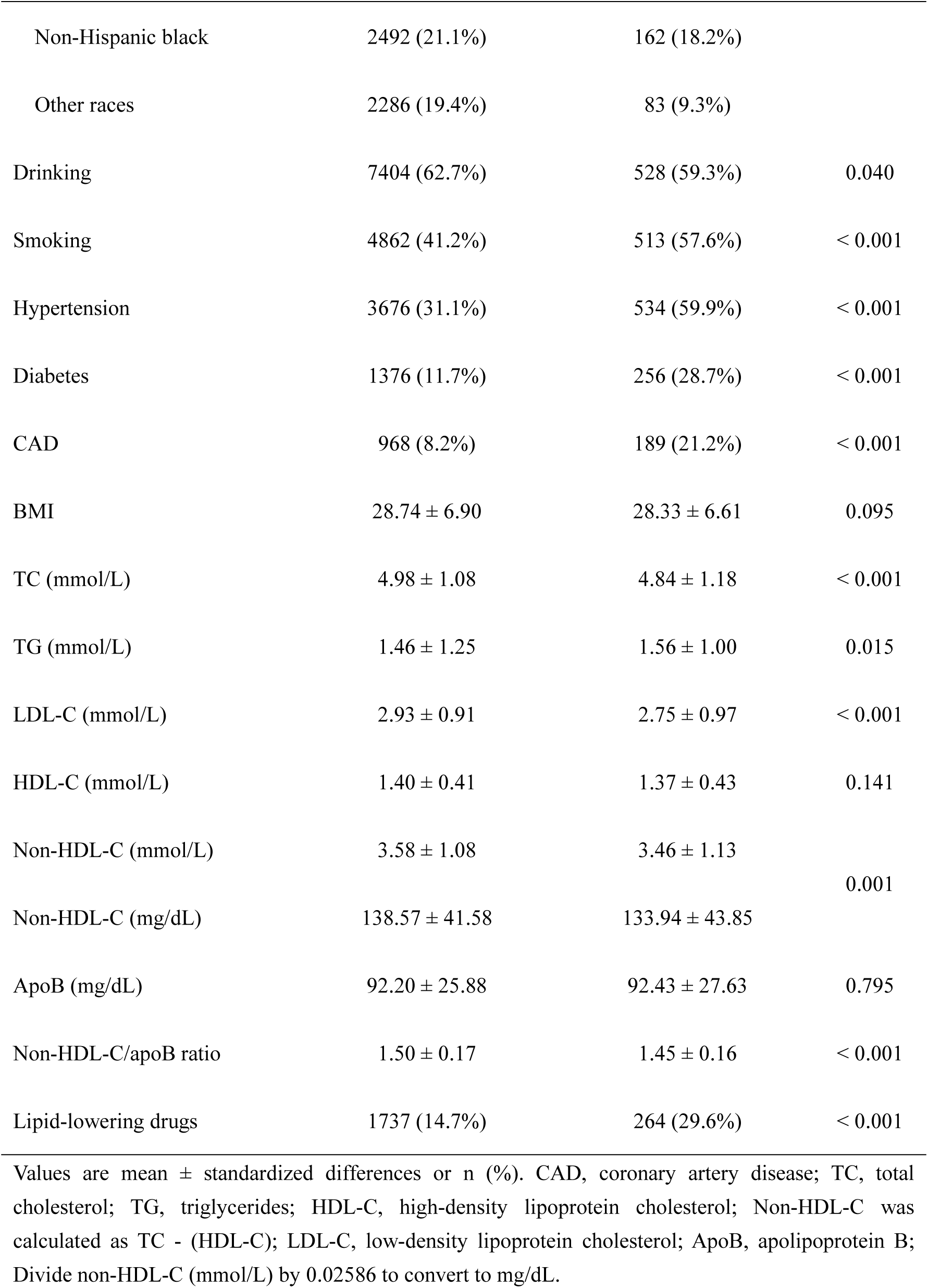
Demographic and baseline characteristics of participants by long-term all-cause mortality.

Based on long-term cardiovascular mortality, the study cohort was categorized into participants without events (n = 12,541) and those with events (n = 156). The demographic and baseline characteristics of the two groups were shown in **Table 2**. Participants who experienced cardiovascular events were older, mainly male, and had a higher prevalence of smoking (all *P* values < 0.001). In the population with cardiovascular mortality, hypertension, diabetes and CAD were more prevalent, as was the use of lipid-lowering drugs (all *P* values < 0.001). Besides, they tended to have higher serum TG and apoB concentrations (both *P* values < 0.01). Importantly, those with cardiovascular events had significantly lower non-HDL-C/apoB ratio (*P* < 0.001).

**Table 2.** Demographic and baseline characteristics of participants by long-term cardiovascular mortality.

|  | No events (n = 12,541) | Events (n = 156) | P value |
| --- | --- | --- | --- |
| Age (years) | 46.54 ± 18.52 | 70.76 ± 12.68 | < 0.001 |
| Age group |  |  |  |
| < 65 years | 10006 (79.8%) | 38 (24.4%) | < 0.001 |
| ≥ 65 years | 2535 (20.2%) | 118 (75.6%) |  |
| Gender |  |  |  |
| male | 6113 (48.7%) | 102 (65.4%) | < 0.001 |
| female | 6428 (51.3%) | 54 (34.6%) |  |
| Race |  |  |  |
| Mexican American | 2139 (17.1%) | 19 (12.2%) | < 0.001 |
| Non-Hispanic white | 5421 (43.2%) | 95 (60.9%) |  |
| Non-Hispanic black | 2625 (20.9%) | 29 (18.6%) |  |
| Other races | 2356 (18.8%) | 13 (8.3%) |  |
| Drinking | 7829 (62.4%) | 103 (66.0%) | 0.356 |
| Smoking | 5281 (42.1%) | 94 (60.3%) | < 0.001 |
| Hypertension | 4110 (32.8%) | 100 (64.1%) | < 0.001 |
| Diabetes | 1584 (12.6%) | 48 (30.8%) | < 0.001 |
| CAD | 1122 (8.9%) | 35 (22.4%) | < 0.001 |
| BMI | 28.72 ± 6.88 | 28.08 ± 6.27 | 0.265 |
| TC (mmol/L) | 4.97 ± 1.09 | 5.01 ± 1.32 | 0.673 |
| TG (mmol/L) | 1.46 ± 1.24 | 1.75 ± 1.26 | 0.004 |
| LDL-C (mmol/L) | 2.92 ± 0.92 | 2.82 ± 1.02 | 0.175 |
| HDL-C (mmol/L) | 1.40 ± 0.41 | 1.35 ± 0.42 | 0.172 |
| Non-HDL-C (mmol/L) | 3.57 ± 1.08 | 3.66 ± 1.32 | 0.344 |
| Non-HDL-C (mg/dL) | 138.21 ± 41.63 | 141.39 ± 51.12 |  |
| ApoB (mg/dL) | 92.14 ± 25.94 | 98.15 ± 30.72 | 0.004 |
| Non-HDL-C/apoB ratio | 1.50 ± 0.17 | 1.43 ± 0.17 | < 0.001 |
| Lipid-lowering drugs | 1943 (15.5%) | 58 (37.2%) | < 0.001 |

Values are mean ± standardized differences or n (%). CAD, coronary artery disease; TC, total cholesterol; TG, triglycerides; HDL-C, high-density lipoprotein cholesterol; Non-HDL-C was calculated as TC - (HDL-C); LDL-C, low-density lipoprotein cholesterol; ApoB, apolipoprotein B; Divide non-HDL-C (mmol/L) by 0.02586 to convert to mg/dL.

### Association of non-HDL-C/apoB ratio with long-term all-cause mortality

The participants were classified into two groups according to whether their non-HDL-C/apoB ratio ≥ or < 1.4 at baseline. K-M survival curves exhibited a significant disparity in the risk of all-cause mortality between the groups (*P* for log-rank test < 0.001, **Figure 2A**). To mitigate effects of confounding variables, potential confounders (age, sex, race, drinking, smoking, hypertension, diabetes, CAD, serum TG level, and lipid-lowering drugs) were incorporated as adjustment factors into multivariate regression. The final Cox regression model revealed a statistically significant association between a decrease in non-HDL-C/apoB ratio and an increased risk of long-term all-cause mortality (HR = 0.51, 95% CI: 0.33-0.80, **Table 3**). The RCS model demonstrated a negative linear correlation between non-HDL-C/apoB ratio and long-term all-cause mortality (*P* for nonlinearity = 0.098, **Figure 3A**).

**Figure 2A.**
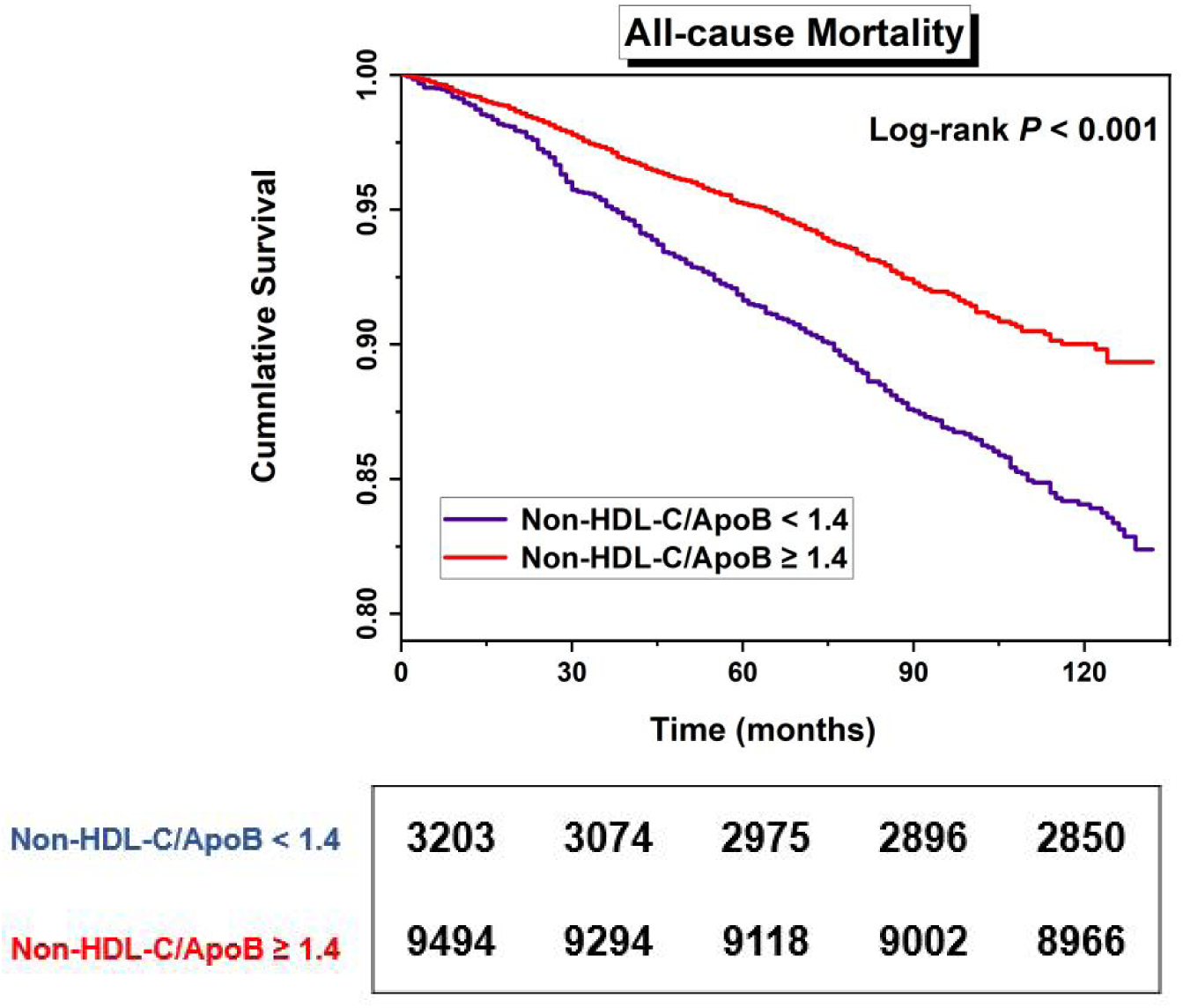
K-M survival analysis for long-term all-cause mortality (Non-HDL-C/apoB ratio < 1.4 *vs.* ≥ 1.4).

**Table 3.**
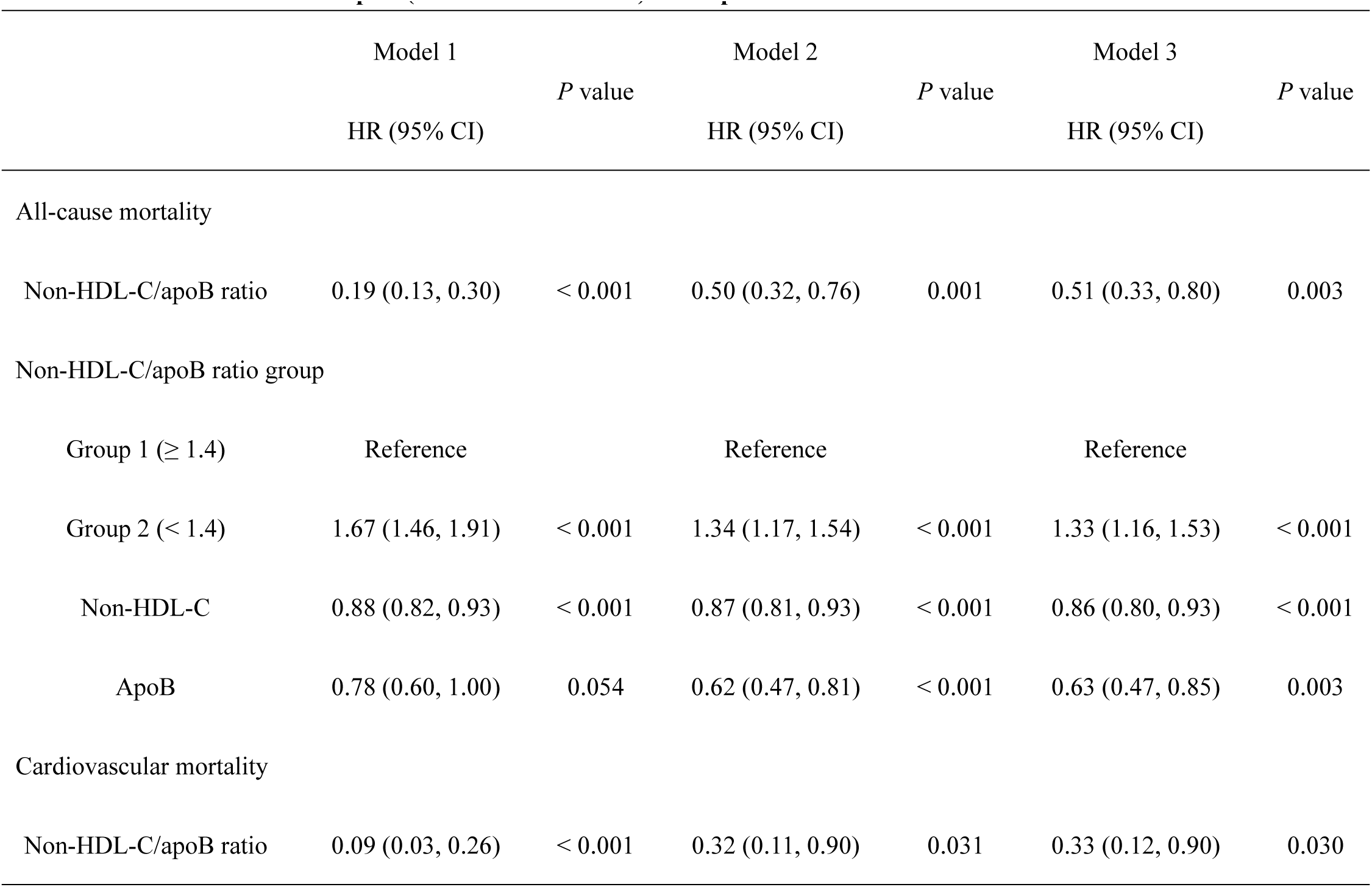
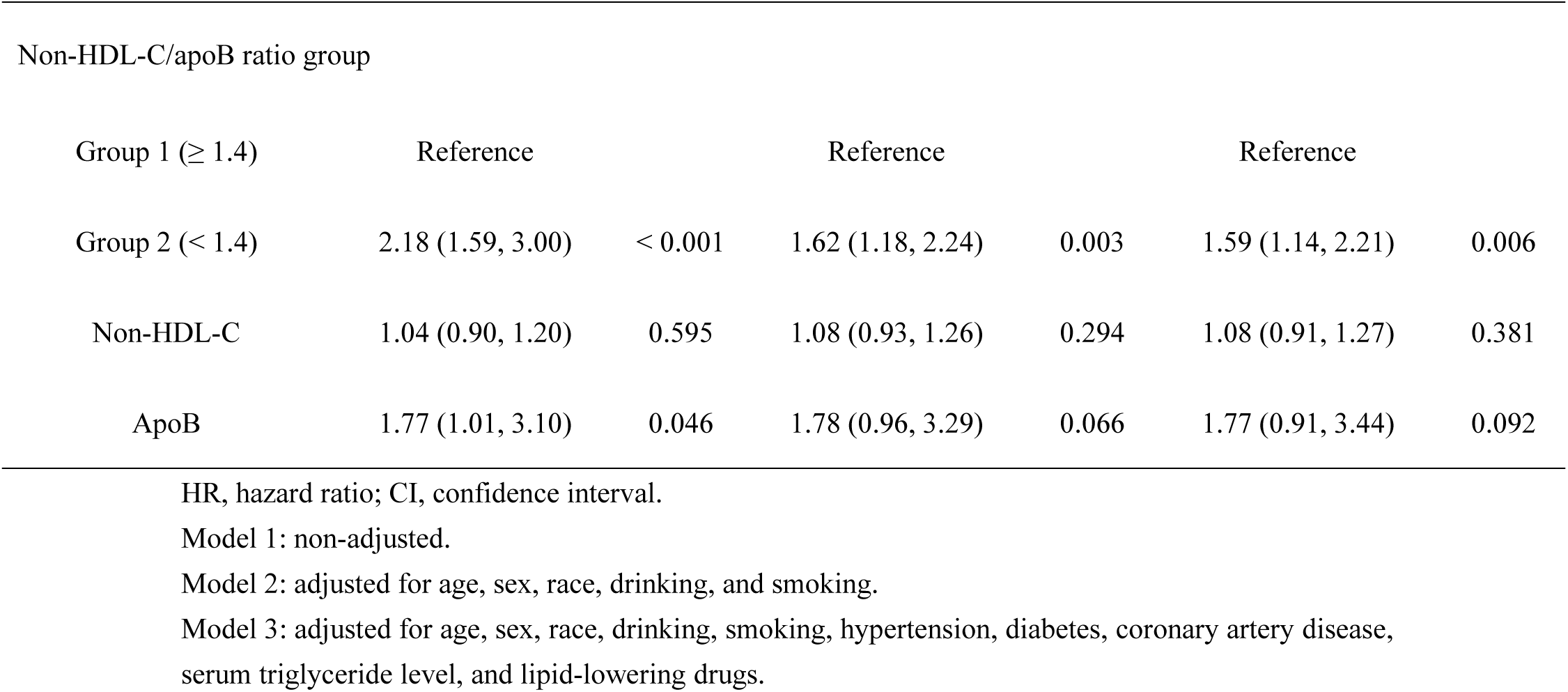
Multivariate Cox models for long-term mortality for non-HDL-C/apoB ratio, non-HDL-C and apoB (continuous variables) in the pooled cohort.

|  | Model 1 |  | Model 2 |  | Model 3 |  |
| --- | --- | --- | --- | --- | --- | --- |
|  | HR (95% CI) | *P* value | HR (95% CI) | *P* value | HR (95% CI) | *P* value |
| All-cause mortality |  |  |  |  |  |  |
| Non-HDL-C/apoB ratio | 0.19 (0.13, 0.30) | < 0.001 | 0.50 (0.32, 0.76) | 0.001 | 0.51 (0.33, 0.80) | 0.003 |
| Non-HDL-C/apoB ratio group |  |  |  |  |  |  |
| Group 1 (≥ 1.4) | Reference |  | Reference |  | Reference |  |
| Group 2 (< 1.4) | 1.67 (1.46, 1.91) | < 0.001 | 1.34 (1.17, 1.54) | < 0.001 | 1.33 (1.16, 1.53) | < 0.001 |
| Non-HDL-C | 0.88 (0.82, 0.93) | < 0.001 | 0.87 (0.81, 0.93) | < 0.001 | 0.86 (0.80, 0.93) | < 0.001 |
| ApoB | 0.78 (0.60, 1.00) | 0.054 | 0.62 (0.47, 0.81) | < 0.001 | 0.63 (0.47, 0.85) | 0.003 |
| Cardiovascular mortality |  |  |  |  |  |  |
| Non-HDL-C/apoB ratio | 0.09 (0.03, 0.26) | < 0.001 | 0.32 (0.11, 0.90) | 0.031 | 0.33 (0.12, 0.90) | 0.030 |
| Non-HDL-C/apoB ratio group |  |  |  |  |  |  |
| Group 1 ($\geq 1.4$) | Reference | | Reference | | Reference | |
| Group 2 ($< 1.4$) | 2.18 (1.59, 3.00) | $< 0.001$ | 1.62 (1.18, 2.24) | 0.003 | 1.59 (1.14, 2.21) | 0.006 |
| Non-HDL-C | 1.04 (0.90, 1.20) | 0.595 | 1.08 (0.93, 1.26) | 0.294 | 1.08 (0.91, 1.27) | 0.381 |
| ApoB | 1.77 (1.01, 3.10) | 0.046 | 1.78 (0.96, 3.29) | 0.066 | 1.77 (0.91, 3.44) | 0.092 |

HR, hazard ratio; CI, confidence interval. Model 1: non-adjusted. Model 2: adjusted for age, sex, race, drinking, and smoking. Model 3: adjusted for age, sex, race, drinking, smoking, hypertension, diabetes, coronary artery disease, serum triglyceride level, and lipid-lowering drugs.

### Association of non-HDL-C/apoB ratio with long-term cardiovascular mortality

The K-M curves showed a statistically significant association between different groups based on non-HDL-C/apoB ratio and the rate of overall survival (*P* for log-rank test < 0.001, **Figure 2B**). The Cox model with full adjustment revealed a substantial association between non-HDL-C/apoB ratio and long-term cardiovascular mortality (HR = 0.33, 95% CI: 0.12-0.90). The low ratio group (non-HDL-C/apoB ratio < 1.4) had a significantly higher risk of long-term cardiovascular mortality, estimated to be 1.59 times greater compared to the high ratio group (HR=1.59, 95% CI: 1.14-2.21, **Table 3**). The RCS analysis presented in **Figure 3B** demonstrated a gradual decline in HRs for long-term cardiovascular mortality as the non-HDL-C/apoB ratio increases (*P* for nonlinearity = 0.314).

**Figure 2B.**
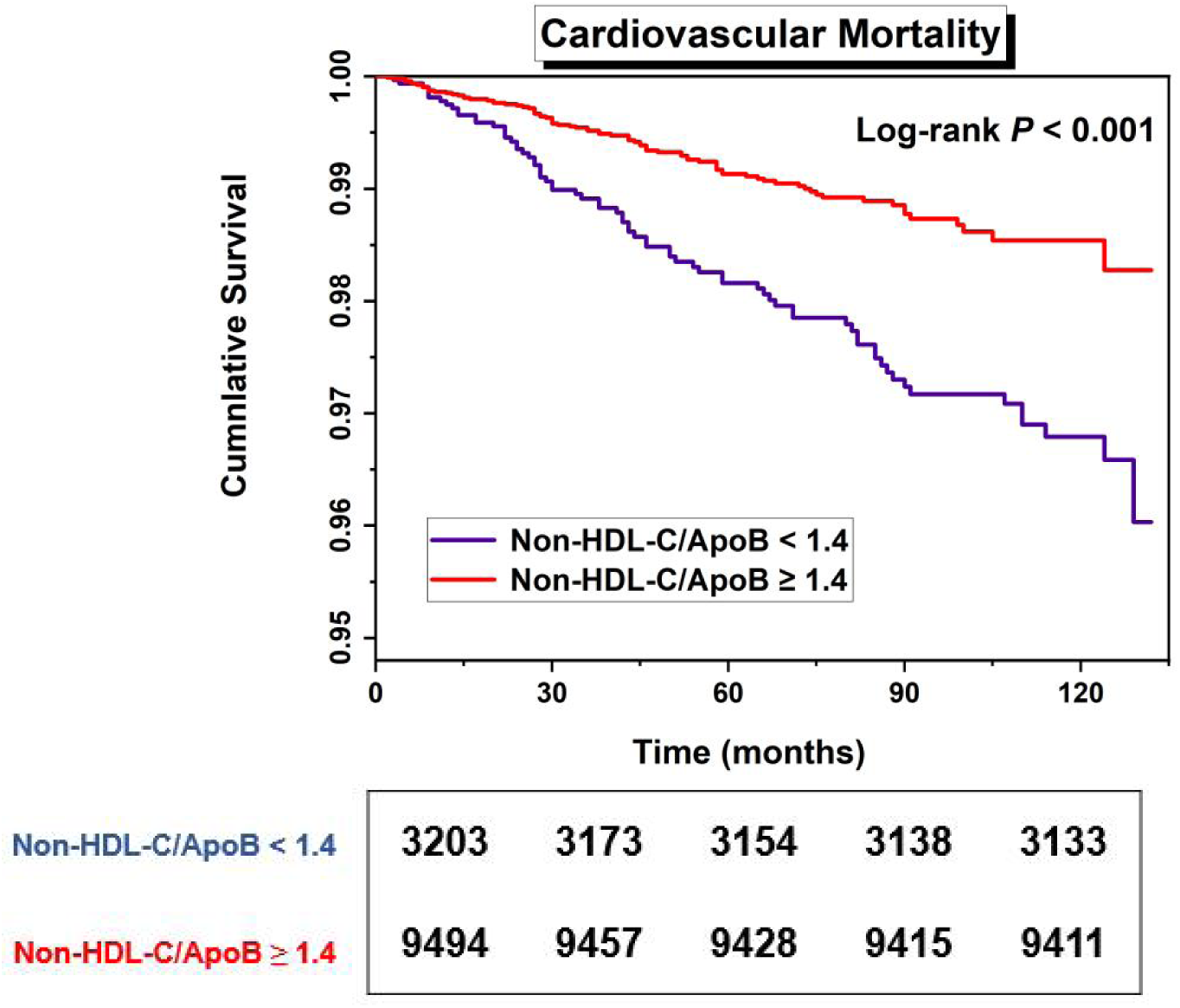
K-M survival analysis for long-term cardiovascular mortality (Non-HDL-C/apoB ratio < 1.4 vs. ≥ 1.4)

**Figure 3.**
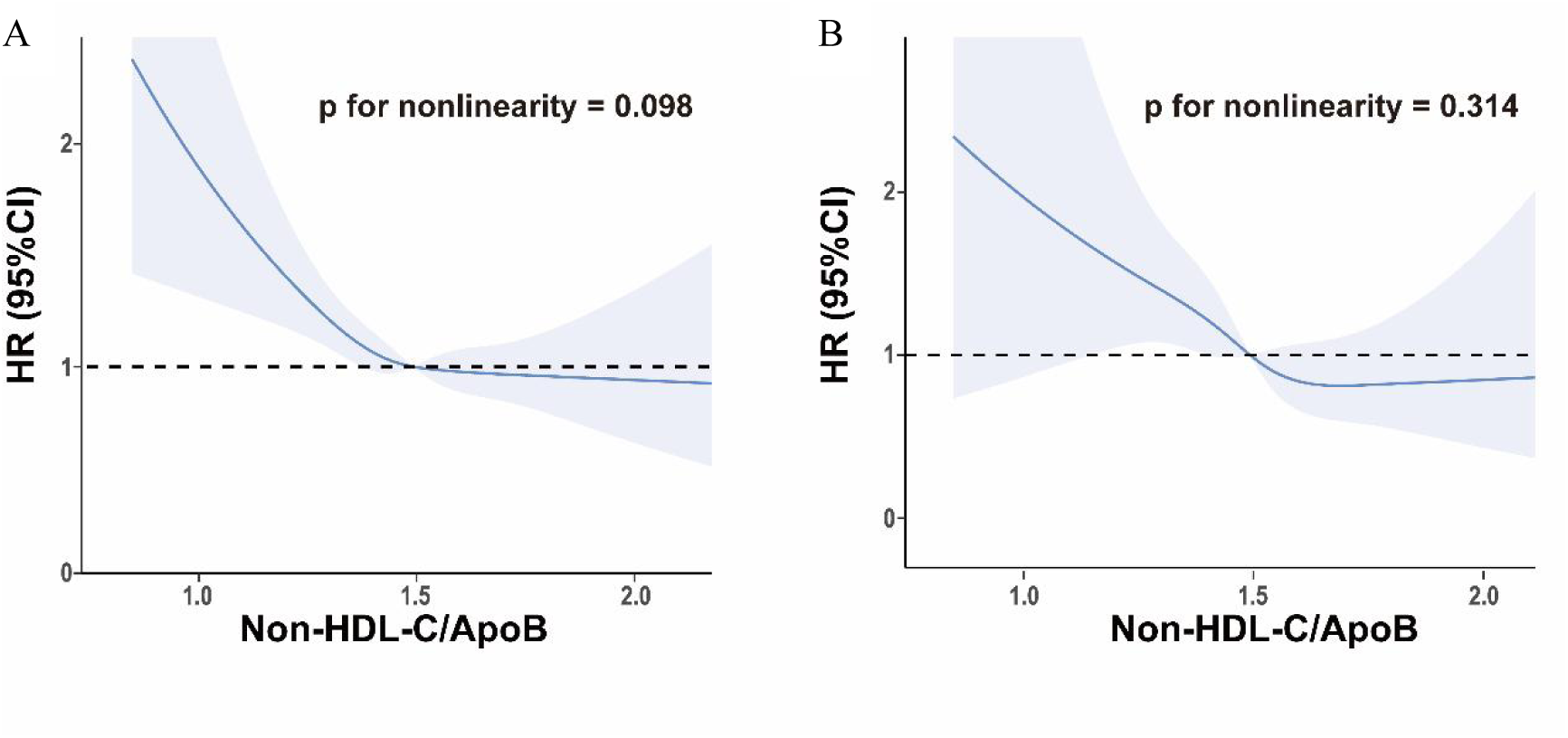
Restricted cubic spline plots of the association between non-HDL-C/apoB ratio with long-term all-cause mortality (A) and cardiovascular mortality (B) in the general population. Analysis was adjusted for age, sex, race, drinking, smoking, hypertension, diabetes, coronary artery disease, serum triglyceride level and lipid-lowering drugs. HR, hazard ratio.

### Subgroup analysis

As presented in **Table 4**, a stratified analysis by age, sex, CAD, diabetes, use of lipid-lowering drugs and hypertriglyceridemia, further explored the association between non-HDL-C/apoB ratio and long-term mortality. There was no statistically significant difference between the non-HDL-C/apoB ratio and long-term mortality among individuals in various age groups (< 65 years and ≥ 65 years), as well as those with and without CAD, diabetes, lipid-lowering medication, and hypertriglyceridemia (all *P* values for interaction > 0.05). Furthermore, a notable interaction was observed between non-HDL-C/apoB ratio and sexes in relation to long-term all-cause and cardiovascular mortality (both *P* values for interaction < 0.05), and the association between non-HDL-C/apoB ratio and mortality was more pronounced in female participants (*P* < 0.01). In exploring of RCS plots to represent non-HDL-C/apoB ratio as a continuous variable, a linear relationship emerged between non-HDL-C/apoB ratio and rates of long-term all-cause and cardiovascular mortality in stratified analysis by age, sex, CAD, diabetes, use of lipid-lowering drugs and hypertriglyceridemia (**Figures 4** and **5**).

**Figure 4.**
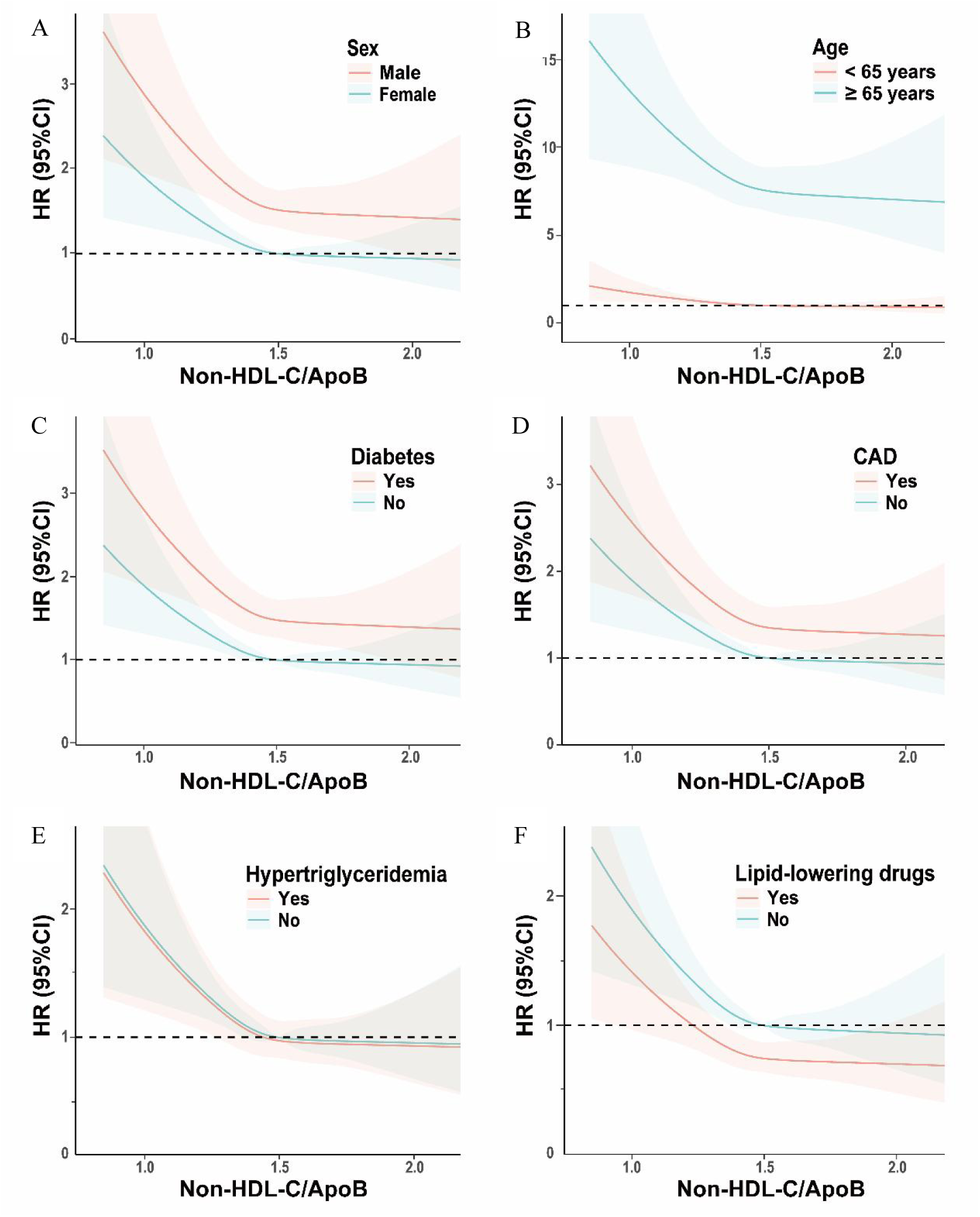
Subgroup analysis of restricted cubic spline plots for the association between non-HDL-C/apoB ratio and long-term all-cause mortality by sex (A), age (B), diabetes (C), coronary artery disease (CAD) (D), hypertriglyceridemia (E) and lipid-lowering drugs (F). Adjusted for age, sex, race, drinking, smoking, hypertension, diabetes, CAD, serum triglyceride level, and lipid-lowering drugs except the subgroup variable.

**Figure 5.**
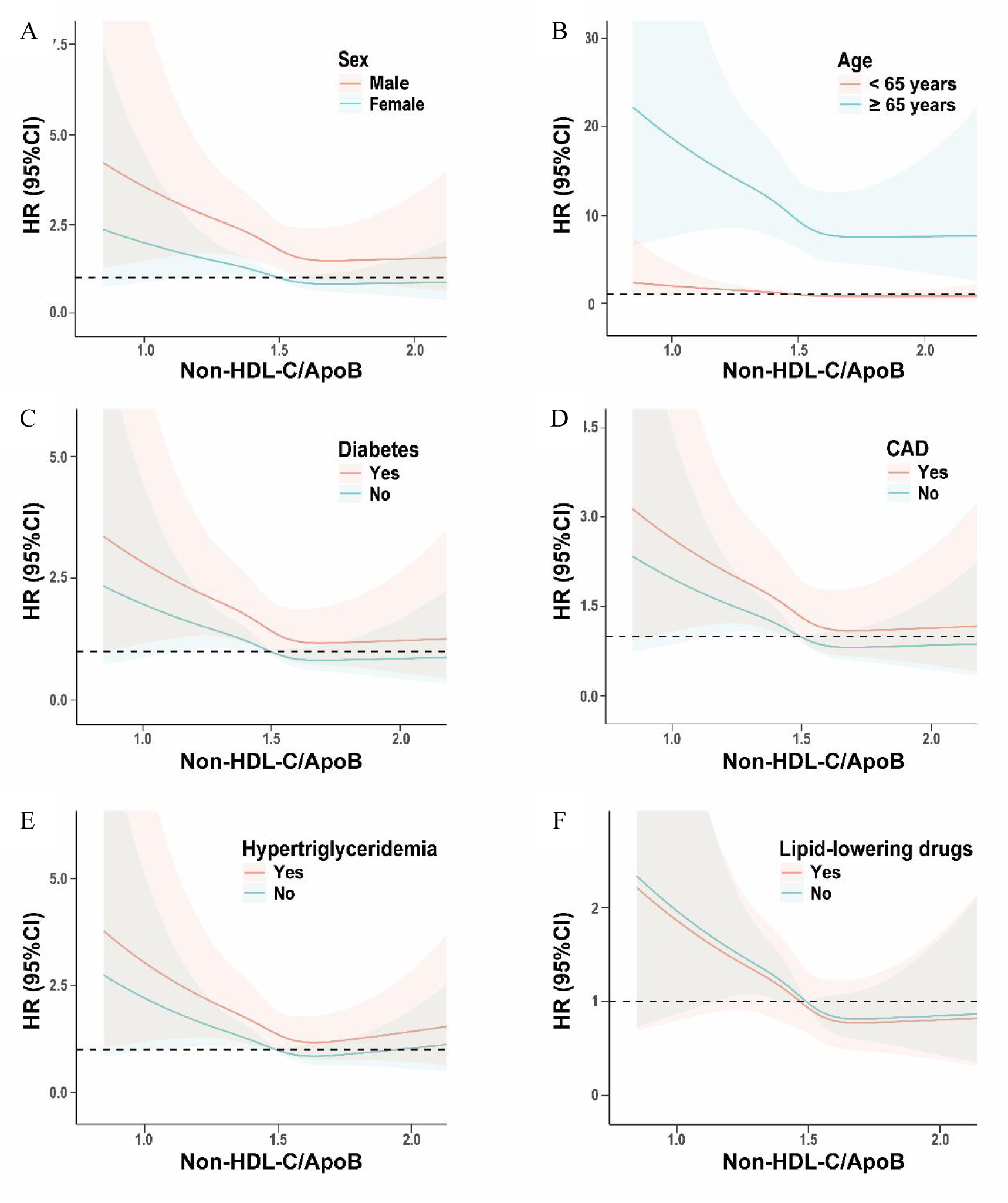
Subgroup analysis of restricted cubic spline plots for the association between non-HDL-C/apoB ratio and long-term cardiovascular mortality by sex (A), age (B), diabetes (C), coronary artery disease (CAD) (D), hypertriglyceridemia (E) and lipid-lowering drugs (F). Adjusted for age, sex, race, drinking, smoking, hypertension, diabetes, CAD, serum triglyceride level, and lipid-lowering drugs except the subgroup variable.

**Table 4.**
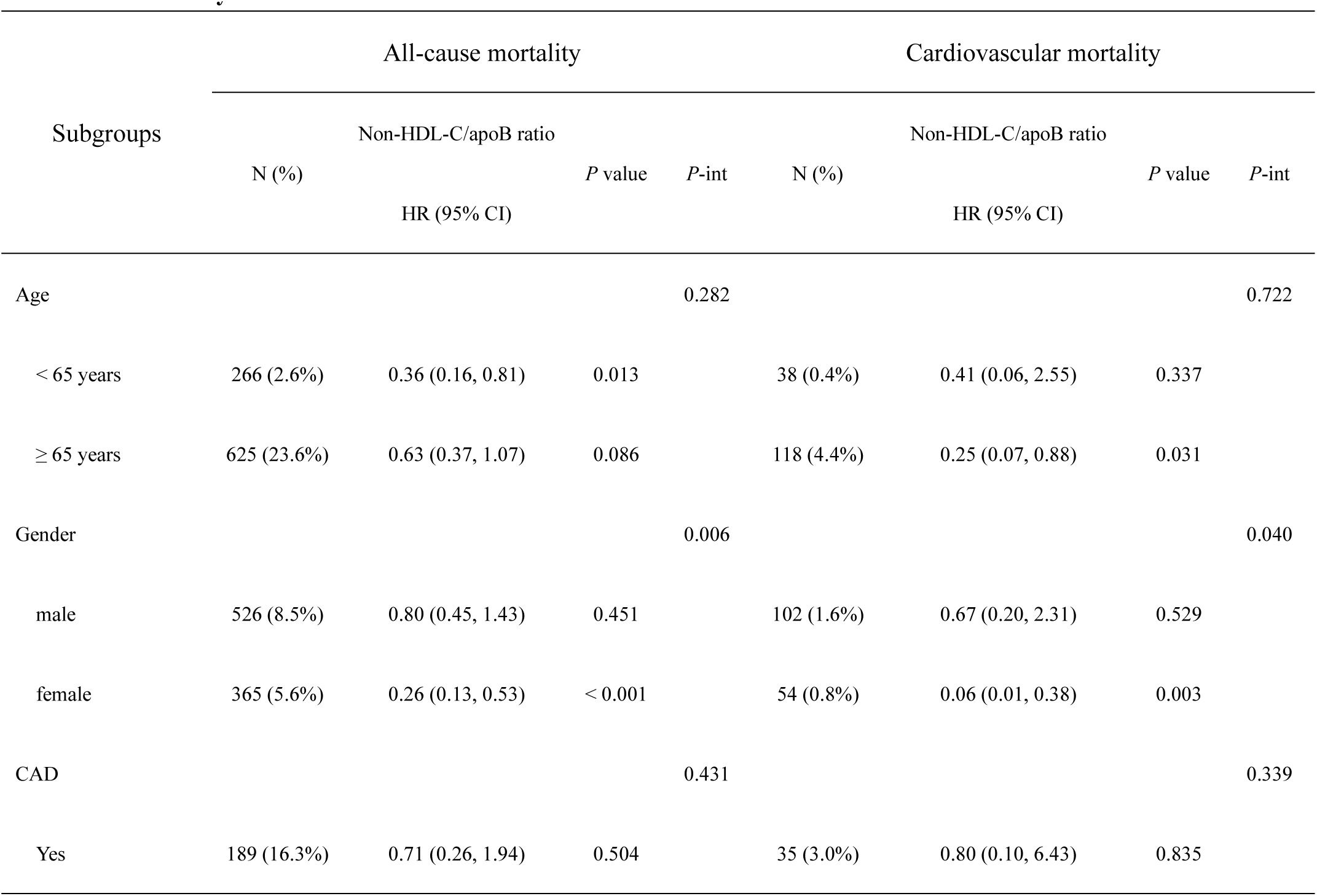
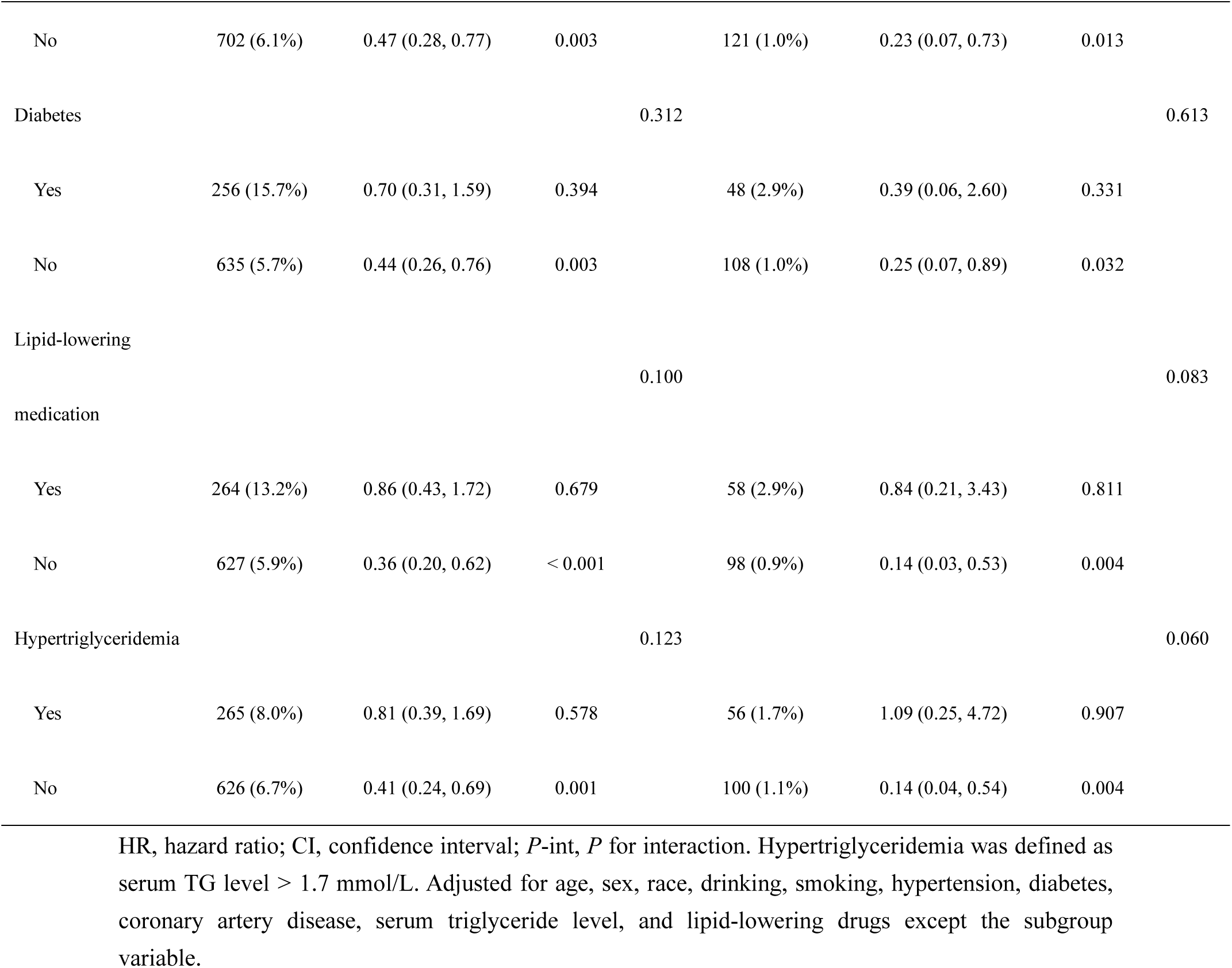
Subgroup analysis for the association between non-HDL-C/apoB ratio and long-term mortality.

## Discussion

To our knowledge, this study is the premiere evaluation exploring the long-term prognostic value of the non-HDL-C/apoB ratio within the general adult population. It indicates that a lower non-HDL-C/apoB ratio was associated with higher long-term mortality, regardless of the presence of other established risk factors. Within the examined population, there was no statistically significant difference observed between non-HDL-C/apoB ratio and the risk of mortality across different age groups, individuals with presence of CAD, diabetes or hypertriglyceridemia, and those with a history of lipid-lowering drugs. Subsequent analysis revealed a negative linear correlation between non-HDL-C/apoB ratio and long-term mortality.

Most guidelines recommend non-HDL-C as the primary target superior to LDL-C for secondary prevention of ASCVD due to its inclusion of additional atherogenic lipoprotein particles, such as very low-density lipoproteins (VLDL), remnants of VLDL, and lipoprotein(a) (Lp(a))[1, 26]. However, non–HDL-C does not differentiate between RC, LDL-C and Lp(a) [7]. When LDL particles are cholesterol-depleted, the concentration of LDL-C decreases, and consequently non-HDL-C will underestimate the risk due to a high number of LDL particles. Besides, the composite marker non-HDL-C is unable to characterize any of dyslipoproteinemias[6]. Furthermore, the treatment targets proposed by guidelines for non-HDL-C are arbitrarily established at a predetermined value of 30 mg/dL above the targets set for LDL-C, which has not been validated[27]. ApoB serves as the structural protein for all non-HDL. It is present in two isoforms—apoB_100_ and apoB_48_[4]. Over 95% of apoB in the circulation is apoB_100_ which binds to non-HDL-C, and apoB_48_ was predominantly detected in chylomicrons and chylomicron remnants particularly in nonfasting specimens. Each atherogenic lipid particle contains a single apoB molecule [4]. Thus, apoB serves as a reliable indicator for quantifying the quantity of atherogenic lipoprotein particles present in serum. In this case, ApoB immunoassays eliminate the necessity of employing lipoprotein subfractionation techniques including NMR spectroscopy to quantify amount of lipoprotein particle.

While apoB is unable to serve as a replacement for NMR-based particle size assessments and cannot distinguish between small and large lipoprotein particles[28], it is currently acknowledged that all non-HDL particles possess identical atherogenicity, irrespective of their size[29]. Based on the above facts, we can make the following conclusions: First, the non-HDL-C concentration cannot truly reflect the number of atherogenic particles. Second, although apoB can provide information about the quantity of atherogenic particles, it does not accurately reflect the precise composition of cholesterol-depleted (TG-enriched) and cholesterol-enriched (TG-depleted) particles. Discordance analysis demonstrates that patients had worse outcomes when atherogenic particle (non-HDL-C or LDL-C) concentrations were lower but the numbers (apoB) were not lower, compared with patients with consistent numbers and concentration[8–10, 30–32]. The prevalence of obesity, diabetes, and hypertension is higher among those with a low non-HDL-C to high apoB ratio[8–10, 30–32]. This inconsistency is easily reflected by the simple physiologically relevant non-HDL-C/apoB ratio. However, these studies on this ratio were intended to illustrate that the measurement of apoB is a more precise marker of cardiovascular risk compared to non-HDL-C. There exists limited research examining the association of non-HDL-C/apoB ratio with long-term mortality. However, it is evident that a low ratio (indicating significant discordance) is associated with elevated cardiovascular risks. This is likely due to an increased concentration of cholesterol-depleted particles (TG-enriched particles) per unit volume compared to other cases. Consequently, non-HDL, when influenced by hypertriglyceridemia, may exhibit increased vulnerability to oxidative modification and dysfunction of HDL, leading to potentially exacerbating the development of atherosclerosis[33–35]. Our study confirms the negative linear association of this ratio with long-term mortality, with a relatively higher risk of death at low ratio (< 1.4). This effect was not observed for non-HDL-C or apoB. First, not all individuals experiencing events (all-cause and cardiovascular deaths) have significantly high levels of these indicators. Furthermore, following the adjustments for age, sex, race, and common confounders related to cardia-cerebrovascular disease, they were not associated with both types of mortality. Therefore, non-HDL-C/apoB ratio is more predictive of long-term mortality than non-HDL-C or apoB alone. Currently, we have no evidence that a ratio of 1.4 serves as a cut-off point between low and high risk of mortality, although in RCS curves it is found that when the ratio exceeds 1.4, there is a notable decrease in the rate of decline in mortality. A study conducted within South Asian individuals has identified that a cut-off value of 1.4 can potentially be employed to evalute the risk of coronary calcification[11]. With regards to the determination of the cut-off value, it is suggested that the study methods pertaining to the LDL-C/apoB ratio be consulted. The classification of small, dense LDL (sdLDL) and large, buoyant LDL is determined by the LDL-C/apoB ratio of 1.2, which is considered a crucial threshold[36–38]. If the ratio < 1.2, it indicates a higher proportion of sdLDL (also cholesterol-depleted), which is a major contributor to ASCVD[36–38]. Mendelian randomization studies indicate a causal relationship between elevated levels of TG-enriched (cholesterol-depleted) lipoproteins or their remnants throughout an individual’s lifespan and an elevated risk of ASCVD and all-cause mortality[39]. The relative fraction of cholesterol-depleted particles in non-HDL-C deserves study due to the inherent complexity in evaluating the relative pathogenicity of various atherogenic lipoprotein particles within non-HDL-C.

The precision of non-HDL-C surpasses that of LDL-C in terms of calculation accuracy due to its independence from TG variability[40] and its ability to be determined reliably regardless of fasting status[41]. However, all laboratory methods for measuring LDL-C, are susceptible to high TG level, leading to inaccurate results[20, 21]. Furthermore, LDL-C was easily determined by the Friedewald formula in NHANES study, which is not recommended by guidelines for patients with TG ≥ 400mg/L[42]. As HDL-C is utilized in the calculations, the presence of analytical errors in the measurement of HDL-C has a significant impact on the computation of non-HDL-C. Although HDL-C test manufacturers strive to comply to the international standardization process, it is important to note that variations in findings may occur due to the diverse methods employed by each manufacturer[43]. Inconsistencies that deviate notably from the ultra-centrifugation reference method are predominantly observed in the circulation of patients presenting with hypertriglyceridemia, mixed dyslipidemia, or other medical conditions that impact the composition and restructuring of lipoproteins. Examples of such conditions include diabetes and chronic renal disease[44]. Despite the measurement of HDL-C in NHANES is a unified precipitation method, this issue needs to be paid attention to in clinical practice. In our study, the results remained significant after adjusting for main factors affecting serum lipids (TG level, diabetes and lipid-lowering medication use).

The current study exhibited some salient aspects and strengths. Initially, it is noteworthy that the sample size was substantial and encompassed a wide range of subjects. The composition of the study cohort closely mirrored that of the broader adult population in the United States. Furthermore, our retrospective cohort study provides robust evidence about the forecasting efficacy of the non-HDL-C/apoB ratio in predicting long-term all-cause and cardiovascular death among individuals in the general population. No prior analogous research has been conducted. At last, we firstly applied RCS models to visualize the correlation between non-HDL-C/apoB ratio and the long-term risk of mortality.

Additionally, there were other potential limits. Primarily, non-HDL-C/apoB ratio was an estimate of amount and relative proportion of cholesterol-depleted particles in all atherogenic lipoprotein particles instead of serving as a direct instrument for differentiating all lipoprotein subtypes. And assessment of non-HDL-C, apoB and other covariates was restricted to the baseline, thus overlooking any changes that may have occurred over the follow-up period. The ratio might incapable of representing the long-term level among adults. In addition, due to incomplete monitoring and thorough analysis of medication adherence and physical activity, it is not possible to rule out the potential impact of lipid-regulating drugs and physical activity on the observed outcomes. Furthermore, data on apoB in NHANES are missing before 2005 and after 2014, and additional data may be required to substantiate our conclusions. Moreover, the data were obtained from individuals residing in the United States, and consequently further investigations could be undertaken in different nations to ascertain the global applicability of these findings.

## Conclusions

In summary, the present study has yielded empirical support for an independent association of non-HDL-C/apoB ratio, serving as an estimation indicator of amounts of cholesterol-depleted particles, with both long-term all-cause and cardiovascular mortality. In contrast to non-HDL-C/apoB ratio ≥ 1.4, the ratio < 1.4 may be associated with higher long-term mortality risk. This ratio could potentially account for some residual risks not explained by non-HDL-C or apoB alone. In the future, it will be imperative to provide evidence elucidating the mechanisms that underlie the association of non-HDL-C/apoB ratio with long-term mortality besides the amount of cholesterol-depleted particles and to clarify the benefits of therapies targeting cholesterol-depleted particles.

## Data Availability

The original contributions presented in the study are included in the article/supplementary material, further inquiries can be directed to the corresponding author/s.

https://www.cdc.gov/nchs/nhanes/

https://www.cdc.gov/nchs/data-linkage/mortality.htm

https://www.n.cdc.gov/nchs/nhanes/analyticguidelines.aspx

## Acknowledgements

We express gratitude to the researchers and participants of the NHANES study for their invaluable contributions.

## Authors’ contributions

ZG and YL designed the study and conceived the paper. KZ and CW performed statistical analysis and drafted the manuscript. YS was responsible for methodology and investigation. LW and ZZ arranged the data and performed visualization. SY and XT contributed to validation. ZG critically revised the manuscript, and all authors read and approved the final manuscript.

## Dear Editor

We note that your journal first reported the association between non-HDL-C/apoB ratio and coronary calcification (Molina CR, Mathur A, Soykan C, Sathe A, Kunhiraman L. Risk Factor Interactions, Non-High-Density Lipoprotein Cholesterol to Apolipoprotein B Ratio, and Severity of Coronary Arteriosclerosis in South Asian Individuals: An Observational Cohort Study. J Am Heart Assoc. 2023 May 16;12(10):e027697. doi: 10.1161/JAHA.122.027697. Epub 2023 May 15. PMID: 37183833; PMCID: PMC10227306), which aroused our great interest. As a result, we further investigated this indicator in the general population.

Due to mailbox restrictions, we are unable to make changes to these authors’ information in the “Author Information” screen. The following authors’ information has been updated (the modifications are highlighted in blue). In addition, please note that Yuan Li was listed as the co-corresponding author to thank him for keeping in touch with all the authors at any time.

Kerui Zhang Center for Cardiovascular Disease, The Affiliated Suzhou Hospital of Nanjing Medical University, Suzhou Municipal Hospital, Gusu School, Nanjing Medical University

Chenchen Wei Center for Cardiovascular Disease, The Affiliated Suzhou Hospital of Nanjing Medical University, Suzhou Municipal Hospital, Gusu School, Nanjing Medical University

Yuan Li Center for Cardiovascular Disease, The Affiliated Suzhou Hospital of Nanjing Medical University, Suzhou Municipal Hospital, Gusu School, Nanjing Medical University

## Reference

1. Mach F, Baigent C, Catapano AL, Koskinas KC, Casula M, Badimon L, Chapman MJ, De Backer GG, Delgado V, Ference BA et al: 2019 ESC/EAS Guidelines for the management of dyslipidaemias: lipid modification to reduce cardiovascular risk. European heart journal 2020, 41(1):111–188.

2. Di Angelantonio E, Gao P, Pennells L, Kaptoge S, Caslake M, Thompson A, Butterworth AS, Sarwar N, Wormser D, Saleheen D et al: Lipid-related markers and cardiovascular disease prediction. Jama 2012, 307(23):2499–2506.

3. Shapiro MD, Fazio S: From Lipids to Inflammation: New Approaches to Reducing Atherosclerotic Risk. Circulation research 2016, 118(4):732–749.

4. Contois JH, McConnell JP, Sethi AA, Csako G, Devaraj S, Hoefner DM, Warnick GR: Apolipoprotein B and cardiovascular disease risk: position statement from the AACC Lipoproteins and Vascular Diseases Division Working Group on Best Practices. Clinical chemistry 2009, 55(3):407–419.

5. Wilkins JT, Li RC, Sniderman A, Chan C, Lloyd-Jones DM: Discordance Between Apolipoprotein B and LDL-Cholesterol in Young Adults Predicts Coronary Artery Calcification: The CARDIA Study. J Am Coll Cardiol 2016, 67(2):193–201.

6. Langlois MR, Nordestgaard BG: Which Lipids Should Be Analyzed for Diagnostic Workup and Follow-up of Patients with Hyperlipidemias? Current cardiology reports 2018, 20(10):88.

7. Nordestgaard BG, Langlois MR, Langsted A, Chapman MJ, Aakre KM, Baum H, Borén J, Bruckert E, Catapano A, Cobbaert C et al: Quantifying atherogenic lipoproteins for lipid-lowering strategies: Consensus-based recommendations from EAS and EFLM. Atherosclerosis 2020, 294:46–61.

8. Sniderman AD, Lamarche B, Contois JH, de Graaf J: Discordance analysis and the Gordian Knot of LDL and non-HDL cholesterol versus apoB. Current opinion in lipidology 2014, 25(6):461–467.

9. Sniderman AD, Islam S, Yusuf S, McQueen MJ: Discordance analysis of apolipoprotein B and non-high density lipoprotein cholesterol as markers of cardiovascular risk in the INTERHEART study. Atherosclerosis 2012, 225(2):444–449.

10. Solnica B, Sniderman AD, Wyszomirski A, Rutkowski M, Chlebus K, Bandosz P, Pencina MJ, Zdrojewski T.: Concordance/discordance between serum apolipoprotein B, low density lipoprotein cholesterol and non-high density lipoprotein cholesterol in NATPOL 2011 participants – An epidemiological perspective. International journal of cardiology 2023, 390:131150.

11. Molina CR, Mathur A, Soykan C, Sathe A, Kunhiraman L: Risk Factor Interactions, Non-High-Density Lipoprotein Cholesterol to Apolipoprotein B Ratio, and Severity of Coronary Arteriosclerosis in South Asian Individuals: An Observational Cohort Study. Journal of the American Heart Association 2023, 12(10):e027697.

12. Blake GJ, Otvos JD, Rifai N, Ridker PM: Low-density lipoprotein particle concentration and size as determined by nuclear magnetic resonance spectroscopy as predictors of cardiovascular disease in women. Circulation 2002, 106(15):1930–1937.

13. Tsai MY, Georgopoulos A, Otvos JD, Ordovas JM, Hanson NQ, Peacock JM, Arnett DK: Comparison of ultracentrifugation and nuclear magnetic resonance spectroscopy in the quantification of triglyceride-rich lipoproteins after an oral fat load. Clinical chemistry 2004, 50(7):1201–1204.

14. Kanonidou C: Small dense low-density lipoprotein: Analytical review. Clinica chimica acta; international journal of clinical chemistry 2021, 520:172–178.

15. Boot CS, Middling E, Allen J, Neely RDG: Evaluation of the Non-HDL Cholesterol to Apolipoprotein B Ratio as a Screening Test for Dysbetalipoproteinemia. Clinical chemistry 2019, 65(2):313–320.

16. Paquette M, Bernard S, Blank D, Paré G, Baass A: A simplified diagnosis algorithm for dysbetalipoproteinemia. Journal of clinical lipidology 2020, 14(4):431–437.

17. Kaneva AM, Potolitsyna NN, Bojko ER: Usefulness of the LDL-C/apoB ratio in the overall evaluation of atherogenicity of lipid profile. Archives of physiology and biochemistry 2017, 123(1):16–22.

18. Drexel H, Larcher B, Mader A, Vonbank A, Heinzle CF, Moser B, Zanolin-Purin D, Saely CH: The LDL-C/ApoB ratio predicts major cardiovascular events in patients with established atherosclerotic cardiovascular disease. Atherosclerosis 2021, 329:44–49.

19. Xiao L, Zhang K, Wang F, Wang M, Huang Q, Wei C, Gou Z: The LDL-C/ApoB ratio predicts cardiovascular and all-cause mortality in the general population. Lipids Health Dis 2023, 22(1):104.

20. Martin SS, Blaha MJ, Elshazly MB, Toth PP, Kwiterovich PO, Blumenthal RS, Jones SR: Comparison of a novel method vs the Friedewald equation for estimating low-density lipoprotein cholesterol levels from the standard lipid profile. Jama 2013, 310(19):2061–2068.

21. Oliveira MJ, van Deventer HE, Bachmann LM, Warnick GR, Nakajima K, Nakamura M, Sakurabayashi I, Kimberly MM, Shamburek RD, Korzun WJ et al: Evaluation of four different equations for calculating LDL-C with eight different direct HDL-C assays. Clinica chimica acta; international journal of clinical chemistry 2013, 423:135–140.

22. Doran B, Guo Y, Xu J, Weintraub H, Mora S, Maron DJ, Bangalore S: Prognostic value of fasting versus nonfasting low-density lipoprotein cholesterol levels on long-term mortality: insight from the National Health and Nutrition Examination Survey III (NHANES-III). Circulation 2014, 130(7):546–553.

23. Singer MV, Feick P, Gerloff A: Alcohol and smoking. Digestive diseases (Basel, Switzerland) 2011, 29(2):177–183.

24. Williams B, Mancia G, Spiering W, Agabiti Rosei E, Azizi M, Burnier M, Clement DL, Coca A, de Simone G, Dominiczak A et al: 2018 ESC/ESH Guidelines for the management of arterial hypertension. European heart journal 2018, 39(33):3021–3104.

25. 2. Classification and Diagnosis of Diabetes: Standards of Medical Care in Diabetes-2020. Diabetes care 2020, 43(Suppl 1):S14–s31.

26. Grundy SM, Stone NJ, Bailey AL, Beam C, Birtcher KK, Blumenthal RS, Braun LT, de Ferranti S, Faiella-Tommasino J, Forman DE et al: 2018 AHA/ACC/AACVPR/AAPA/ABC/ACPM/ADA/AGS/APhA/ASPC/NLA/PCNA Guideline on the Management of Blood Cholesterol: A Report of the American College of Cardiology/American Heart Association Task Force on Clinical Practice Guidelines. Circulation 2019, 139(25):e1082–e1143.

27. Elshazly MB, Martin SS, Blaha MJ, Joshi PH, Toth PP, McEvoy JW, Al-Hijji MA, Kulkarni KR, Kwiterovich PO, Blumenthal RS et al: Non-high-density lipoprotein cholesterol, guideline targets, and population percentiles for secondary prevention in 1.3 million adults: the VLDL-2 study (very large database of lipids). J Am Coll Cardiol 2013, 62(21):1960–1965.

28. Cole TG, Contois JH, Csako G, McConnell JP, Remaley AT, Devaraj S, Hoefner DM, Mallory T, Sethi AA, Warnick GR: Association of apolipoprotein B and nuclear magnetic resonance spectroscopy-derived LDL particle number with outcomes in 25 clinical studies: assessment by the AACC Lipoprotein and Vascular Diseases Division Working Group on Best Practices. Clinical chemistry 2013, 59(5):752–770.

29. Sniderman AD, Thanassoulis G, Glavinovic T, Navar AM, Pencina M, Catapano A, Ference BA: Apolipoprotein B Particles and Cardiovascular Disease: A Narrative Review. JAMA cardiology 2019, 4(12):1287–1295.

30. Cromwell WC, Otvos JD, Keyes MJ, Pencina MJ, Sullivan L, Vasan RS, Wilson PW, D’Agostino RB: LDL Particle Number and Risk of Future Cardiovascular Disease in the Framingham Offspring Study – Implications for LDL Management. Journal of clinical lipidology 2007, 1(6):583–592.

31. Mora S, Buring JE, Ridker PM: Discordance of low-density lipoprotein (LDL) cholesterol with alternative LDL-related measures and future coronary events. Circulation 2014, 129(5):553–561.

32. Otvos JD, Mora S, Shalaurova I, Greenland P, Mackey RH, Goff DC, Jr.: Clinical implications of discordance between low-density lipoprotein cholesterol and particle number. Journal of clinical lipidology 2011, 5(2):105–113.

33. Tani S, Yagi T, Atsumi W, Kawauchi K, Matsuo R, Hirayama A: Relation between low-density lipoprotein cholesterol/apolipoprotein B ratio and triglyceride-rich lipoproteins in patients with coronary artery disease and type 2 diabetes mellitus: a cross-sectional study. Cardiovascular diabetology 2017, 16(1):123.

34. Skeggs JW, Morton RE: LDL and HDL enriched in triglyceride promote abnormal cholesterol transport. Journal of lipid research 2002, 43(8):1264–1274.

35. Kwiterovich PO, Jr.: Clinical relevance of the biochemical, metabolic, and genetic factors that influence low-density lipoprotein heterogeneity. The American journal of cardiology 2002, 90(8a):30i–47i.

36. Arai H, Kokubo Y, Watanabe M, Sawamura T, Ito Y, Minagawa A, Okamura T, Miyamato Y: Small dense low-density lipoproteins cholesterol can predict incident cardiovascular disease in an urban Japanese cohort: the Suita study. Journal of atherosclerosis and thrombosis 2013, 20(2):195–203.

37. Jin JL, Zhang HW, Cao YX, Liu HH, Hua Q, Li YF, Zhang Y, Wu NQ, Zhu CG, Xu RX et al: Association of small dense low-density lipoprotein with cardiovascular outcome in patients with coronary artery disease and diabetes: a prospective, observational cohort study. Cardiovascular diabetology 2020, 19(1):45.

38. Ikezaki H, Lim E, Cupples LA, Liu CT, Asztalos BF, Schaefer EJ: Small Dense Low-Density Lipoprotein Cholesterol Is the Most Atherogenic Lipoprotein Parameter in the Prospective Framingham Offspring Study. Journal of the American Heart Association 2021, 10(5):e019140.

39. Nordestgaard BG: Triglyceride-Rich Lipoproteins and Atherosclerotic Cardiovascular Disease: New Insights From Epidemiology, Genetics, and Biology. Circulation research 2016, 118(4):547–563.

40. Langlois MR, Nordestgaard BG, Langsted A, Chapman MJ, Aakre KM, Baum H, Borén J, Bruckert E, Catapano A, Cobbaert C et al: Quantifying atherogenic lipoproteins for lipid-lowering strategies: consensus-based recommendations from EAS and EFLM. Clinical chemistry and laboratory medicine 2020, 58(4):496–517.

41. Nordestgaard BG, Langsted A, Mora S, Kolovou G, Baum H, Bruckert E, Watts GF, Sypniewska G, Wiklund O, Borén J et al: Fasting is not routinely required for determination of a lipid profile: clinical and laboratory implications including flagging at desirable concentration cut-points-a joint consensus statement from the European Atherosclerosis Society and European Federation of Clinical Chemistry and Laboratory Medicine. European heart journal 2016, 37(25):1944–1958.

42. Wilson PWF, Jacobson TA, Martin SS, Jackson EJ, Le NA, Davidson MH, Vesper HW, Frikke-Schmidt R, Ballantyne CM, Remaley AT: Lipid measurements in the management of cardiovascular diseases: Practical recommendations a scientific statement from the national lipid association writing group. Journal of clinical lipidology 2021, 15(5):629–648.

43. Langlois MR, Chapman MJ, Cobbaert C, Mora S, Remaley AT, Ros E, Watts GF, Borén J, Baum H, Bruckert E et al: Quantifying Atherogenic Lipoproteins: Current and Future Challenges in the Era of Personalized Medicine and Very Low Concentrations of LDL Cholesterol. A Consensus Statement from EAS and EFLM. Clinical chemistry 2018, 64(7):1006–1033.

44. Miller WG, Myers GL, Sakurabayashi I, Bachmann LM, Caudill SP, Dziekonski A, Edwards S, Kimberly MM, Korzun WJ, Leary ET et al: Seven direct methods for measuring HDL and LDL cholesterol compared with ultracentrifugation reference measurement procedures. Clinical chemistry 2010, 56(6):977–986.

